# How do socio-demographic status and personal attributes influence adherence to COVID-19 preventive behaviours?

**DOI:** 10.1101/2020.08.21.20179663

**Authors:** Shahadat Uddin, Tasadduq Imam, Matloob Khushi, Arif Khan, Mohammad Ali Moni

## Abstract

This study assesses how socio-demographic status and personal attributes influence protection behaviours during a pandemic, with protection behaviours being assessed through three perspectives – social distancing, personal protection behaviour and social responsibility awareness. The COVID-19 preventive behaviours were explored and compared based on the social-demographic and personal attributes of individuals. Using a publicly available and recently collected dataset on Japanese citizens during the COVID-19 early outbreak and exploiting both Classification and Regression Tree (CART) and regression analysis, the study notes that socio-demographic and personal attributes of individuals indeed shape the subjective prevention actions and thereby the control of the spread of a pandemic. Three socio-demographic attributes – sex, marital family status and having children – appear to have played an influential role in Japanese citizens’ abiding by the COVID-19 protection behaviours, especially with women having children being noted more conscious than the male counterparts. Work status also appears to have some impact especially concerning social distancing and personal protection behaviour. Among the personality attributes, smoking behaviour appeared as a contributing factor with non-smokers or less-frequent smokers more compliant to the protection behaviours. Overall, the findings imply the need of public policy campaigning to account for variations in protection behaviour due to socio-demographic and personal attributes during a pandemic.

## 1. Introduction

There have been several outbreaks of coronavirus in the past, namely SARS (severe acute respiratory syndrome) and MERS (Middle East respiratory syndrome) [1]. The latest novel strain appeared in Wuhan, China in 2019 and was named COVID-19 by the World Health Organization (WHO). COVID-19 infection could be asymptomatic or could show mild to severe pneumonia like symptoms [2]. The commonly noted symptoms include fever, dry cough, tiredness, less common symptoms include headache, chest pain, loss of speech, difficulty in breathing, sore throat, diarrhoea, conjunctivitis or other aches and pains [3]. A noticeable feature of the disease is its highly contagious nature and, while the virus outbreak is still under investigation, there are views that the spread can happen person-to-person due to close contacts, airborne particles, and contact with contaminated surfaces [4]. Additionally, the high risk group of the COVID-19 is different from recent pandemics with the current strain particularly deadly for older individuals (Tang et al., 2020). Thus, while the world has confronted multiple epidemics in recent times and different countries have had several preparedness plans in place following their past experiences (Henry, 2019; Itzwerth et al., 2018; Jennings, 2009), the current pandemic has come as an unprecedented context in many countries. Indeed, even after different public health measures instituted by the governments, the virus has continued to spread across countries. As of 1^st^ June 2020, WHO Situation Report identifies over 6 million positive cases globally and close to 4 hundred thousand deaths from the disease [5].

Despite the uniqueness of the COVID-19 context, there have been multiple commonly adopted public health measures and recommendations across countries. These include staying at home, social distancing, contact tracing, exercising, and wearing masks, washing hands more often, wearing gloves and using hand sanitiser. There have also been public and media communications to increase public awareness. However, as experiences from the recent pandemic shows, individual behaviour during an epidemic can be influenced by various factors including trust about the Government’s advice and subjective perceptions (Freimuth et al., 2014; Teasdale et al., 2012). Additionally, research notes that individual health protection behaviour during an influenza pandemic can be characterised by social contexts (Chuang et al., 2015).

Thus, with many COVID-19 cases being asymptotic, leading to people carrying on with their daily life as normal and thereby acting as virus carriers and threat especially to elderly people, it is interesting to explore the public behaviour and their understanding of the risk associated with COVID-19 spread. This can guide policy makers on adjusting measures to reduce the risk of further outbreaks. In this study, we analyse data from Japan to understand on what level people implemented these guidelines and the impacts of socio-demographic status and personal attributes had on their behaviour. Further, while existing studies on pandemics have generally utilised traditional statistical approaches, we apply in this research the Classification and Regression Tree (CART), a machine learning algorithm, for a robust analysis.

## 2. Literature Review

Implementing large scale lockdown and social distancing was difficult for high populous nations such as Indonesia, however, modelling shows early interventions have saved thousands of lives [6]. A WeChat survey of 3083 participants concludes that people living in cities have a better understanding of the disease compared to those in rural areas [7]. The study also demonstrates males (OR: 0.544, 95% CI: 0.440–0.673), younger adults (1.844, 1.466–2.320), and subjects with higher education (2.200, 1.780–2.718) exhibited better behaviours when protective measures such as washing hands, wearing masks or exercising were given [7]. This finding indicates that health education should be strengthened among adults living in rural areas where access to medical services is limited. A survey of 976 university academics and students of the Iraqi Kurdistan region revealed that the perceived risk of contracting COVID-19 infection, serious illness and death (26.9%, 29.7%, and 41.7%, respectively) is low in the participants [8]. Similarly, a Bangladeshi cross sectional study of 320 adults identified people living in the urban area (p< 0.01), high education (p< 0.01), rich (p< 0.01) and joint family (p< 0.01) had the most contributions to good practice [9].

The reception of general public plays a key role in controlling an infectious disease such as COVID-19. The best example of this is Singapore where the disease was very well controlled compared to other countries. The key factors of this success come from the experience that the Singapore for a robust analysis. government and its public have experienced during their outbreaks. In 2000, 2003 and 2009 they experienced hand-foot-mouth disease, SARS and H1N1 outbreaks respectively [10]. They learnt that response to such a pandemic has to be well-coordinated and multi-stakeholder approach including participation from public.

The issues that were caused by COVID-19 are unparalleled and unprecedented. This generation has not seen anything like it. It has been argued that the road to full recovery will be very long and multiple waves of infection are predicted [11]. Hence governments face myriad of multi-dimensional challenges including surge in demand for public health system, economical and mental issues of the public. Therefore, this study analyses the effect of personal attributes, age, social and demographics on public behaviour and their perception about the spread of the disease and means to control it.

## 3. Materials

### 3.1 Data source

This study used an open-source research dataset that was made available by Yamamoto [12]. This dataset consists of the survey responses of 11,342 Japanese who were recruited through a cross-sectional approach from a pool of 1.2 million registered individuals. A non-probabilistic quota sampling was followed in this survey so that the sample distributions among gender (male or female), age group (20s, 30s, 40s, 50s or 60s) and employment status (regular employee, non-regular employee, self-employed and not working) become the representative ones compared with their respective national statistics [12].

### 3.2 Variable quantification

From the research dataset, this study considered one question to quantify its three dependent variables - “*Have you ever conducted anything to prevent novel coronavirus infections or outbreaks?*” [12]. For the response of this question, each respondent needs to select one of the five options (‘*Very true*’, ‘*True*’, ‘*Neither*’, ‘*Not true*’ and ‘*Not at all*’) against each of the 21 activities. A very recently published article on the same dataset focused on the same question to characterise the Japanese citizens’ behaviour across three government recommended measures [13]. We consider the same question and items are labelled exactly as in the research dataset [12, 13]. However, our approach in assessment of pandemic protection behaviour is different. The published work examines the citizens’ behaviour and attitudes through a common umbrella of social distancing and personal etiquettes and focuses mainly on 12 items separately within the same question [13]. We, by contrast, utilise all 21 items and group these activities across three customised pandemic protection behaviours – ‘*social distancing*’, ‘*personal protection behaviour*’ and ‘*social responsibility behaviour*’.

Table 1 details the 21 activities grouped across the three dependent variables. More precisely, rather than considering the items in general, we interpret the meaning and impacts of the respective behaviour to form the grouping. For example, stockpiling masks or food can be interpreted as socially irresponsible behaviour especially if the general populace misses out on essential goods due to mass buying by some individuals. While eating nutritious diet or getting rest and exercise may be interpreted as personal behaviour, we consider the social side of the impacts of such behaviour and the need for a healthy society – thus, these items are also grouped under social responsibility. Similarly, behaviours like avoiding closed but ill-ventilated spaces, not speaking in close proximity, avoiding mass gathering, and related practices are grouped under social distancing behaviours; while taking measures to disinfect surroundings, wearing masks, avoiding travels for any reason when having cold, and related practices are deemed as personal protection behaviours.

**Table 1:** List of activities used for different dependent variables of this study.

| **Social distancing** | **Personal protection behaviour** | **Social responsibility awareness** |
| --- | --- | --- |
| 1. Avoid a poorly ventilated closed space | 1. Undertake frequent handwashing | 1. Stockpile surgical-style mask |
| 2. Avoid large gatherings | 2. Undertake cough etiquette | 2. Stockpile food, toilet paper, tissue paper, etc. |
| 3. Avoid conversations or shouting in proximity | 3. Disinfect things around | 3. Avoid contact with younger people |
| 4. Avoid places where items 1-3 above overlap | 4. Avoid going out when you have a cold | 4. Avoid contact with the older people |
| 5. Do not go to dinner with friends | 5. Avoid going to clinic even when having a cold symptom | 5. Get enough rest and sleep |
| 6. Do not go to mass gatherings | 6. Prepare consultation and transportation methods for when you feel ill | 6. Eat a nutritious diet |
| 7. Participate in virtual events using online tools | 7. Always wear a surgical-style mask when going out | 7. Do exercise such as a jogging or exercise using DVD |

This study considers two broad categories of independent variables – personal attributes and socio-demographic attributes. For the personal attribute category, three variables are used: (i) Having children younger than junior high school age; (ii) Smoking frequency; and (iii) Drinking frequency. The first one has a binary response (yes/no). Smoking frequency has four choices (‘*Every day*’, ‘*Sometimes*’, ‘*Used to smoke but do not now*’ and ‘*Never smoked*’); whereas, drinking frequency offers six choices (‘*Never drink*’, ‘*I used to drink but I quitted*’, ‘*Few times per month*’, ‘*1–2 times per month*’, ‘*3–6 times per month*’ and ‘*Every day*’). Six socio-demographic variables are considered in this study. They are – (i) Age (with five choices, 20s, 30s, 40s, 50s and 60–64); (ii) Gender (female/male); (iii) Marital status (Married or Not married); (iv) Education (university/college graduate or not); (v) Work status (four choices – ‘*Regular employee*’, ‘*Non-regular employee*’, ‘*Self-employed or others*’ and ‘*Not working*’); and (vi) Household annual income (10 ascending choices – ‘*less than 2,000K Japanese yen*’, seven choices between 2,000K and 19,999K Japanese yen, ‘*More than 20,000K Japanese yen*’ and ‘*Do not know*’). The present literature has guided us, as outlined in the literature review section, in selecting these personal and socio-demographic attributes.

### 3.3 Data analysis framework

Figure 1 shows the data analysis framework followed in this research. To convert the categorical responses into a numerical one, the RIDIT analysis has been applied to the dependent variables of this research. After this, CART is applied to identify important independent variables. Finally, regression is used to model the dependent variables using the independent variables. Here is a brief about each of these three methods.

**Figure 1:**
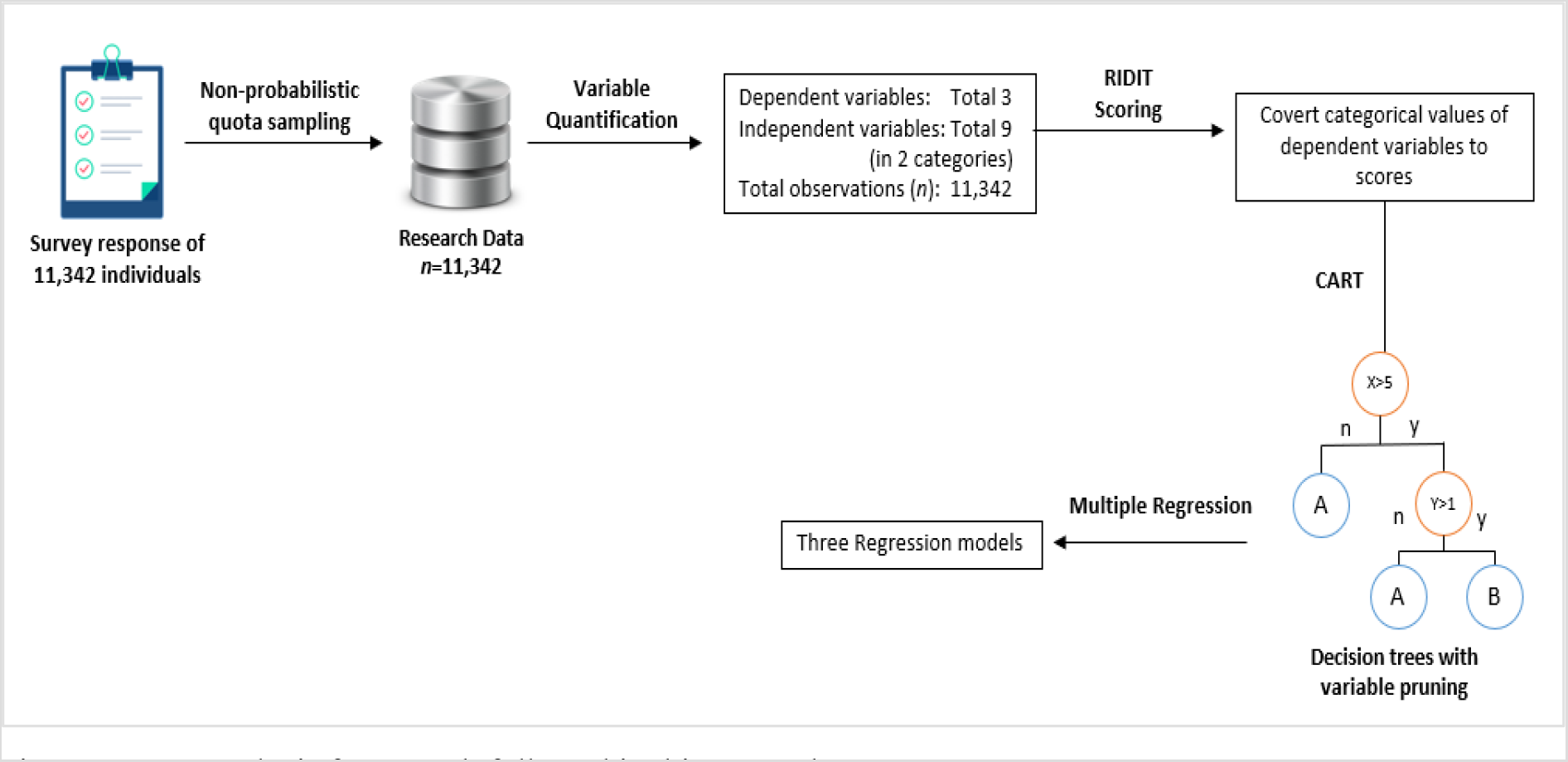
Data analysis framework followed in this research

#### 3.3.1 RIDIT scoring

RIDIT is a statistical method for analysing subjectively categorized response variable, i.e., ordered qualitative measurements often called as ordinal data [14, 15]. For example, if the response variable has subjective scales with ordered categories, such as – *“never smoked”, “smokes sometimes”, “smokes every day”* or *“minor pain”, “moderate pain”, “severe pain”* etc., then they may not be adequately analysed by chi-square- or t-test-based statistical methods. RIDIT analysis is often used to transform the response variable by allocating scores relative to the identified (or reference) distribution of the data.

The term RIDIT stands for “*relative to an identified distribution integral transformation*” and this was termed by Bross for its analogy with other statistical transformation methods such as *probit* and *logit* [16]. Essentially, the process of RIFIT analysis can be summarised in the following steps:

*Step 1:* Select the categorical response variable whose response values are categorical, subjective and in order. For example, for the survey question – “how often do you smoke”, the response values are – “never”, “sometimes” and “daily”. Respondents giving the same response is called a group.
*Step 2:* Choose references or identified distribution data. Based on this set, the non-reference data will be baselined. For this reference set, each group is assigned a score or weight. Thus, each of the response values or categories are transformed into a score. Let us assume, a response variable *X* has *n* ordered responses in the reference set marked as – *x*_1_, *x*_2_, *x*_3_,…, *x*_*n*_. *n*_*i*_ denotes the total number of responses for *x*_*i*_ (*n* = *n*_1_ + *n*_2_ + ⋯ + *n*_*i*_ + ⋯ + *n*_*n*_). Then, RIDIT *R*_*i*_ for response *x*_*i*_ will be,

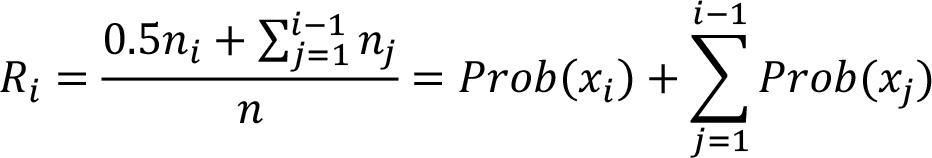 Here *Prob*(*x_i_*) is the probability of choosing response *x*_*i*_ in the reference set.
*Step 3:* The score is assigned to each group in the non-reference or test set and a mean RIDIT score is calculated for each group. The score relates to the probability that a member of the group differs (i.e., have higher score) from a member of the reference population [14]. For this research, we used RIDIT scores as described in step 2 above to transform the categorical responses considered for three dependent variables (drinking and smoking behaviour, household annual income range) into numerical scores. The reference set consisted of the 11,342 respondents who were sampled from the whole dataset through non-probabilistic quota sampling.

#### 3.3.2 CART

CART (short for Classification and Regression Trees) is an umbrella term [17] for referring to two types of decision trees – “classification tree” and “regression tree”. *Decision tree* or *decision tree learning* is a widely used predictive modelling approach used in machine learning, data mining and statistics. Given a dataset with multiple observation of an outcome variable with respect to one or more input (i.e., independent) variable, the decision tree essentially generates a binary tree to predict the outcome variable based on the input variable. The binary tree starts at the single root node and keeps splitting in every non-leaf node into two branches based on a logical comparison of an input variable, e.g., “if income > 800$: Yes/No”, “if sex is male: Yes/No”, etc. The leaf nodes indicate an outcome as the prediction result of the decision tree. Thus, the outcome of a set of input variable values (of future observation) can be predicted by simply following the decision tree logic and reaching towards a particular leaf-node. Now, if the outcome variable has discrete values, i.e., discrete classes such as sex (male, female), disease outcome (benign, malignant), etc., then it is a classification tree. And, if the outcome variable is a continuous variable (price, age in years etc.) then it is a regression tree. CART is a modern term that encompasses a range of algorithms to generate both types of trees.

For the CART implementation in our research, we generated three CART trees (regression trees as the outcome is a continuous number) based on three outcome variables – social distance, personal protection behaviour, social responsibility awareness. In each case, nine independent or input variables are used. Now, when CART generates the decision tree, we applied pruning on the tree to reduce the number of input variables and subsequently reduce the number of splitting in the tree to generate a smaller tree and avoid overfitting of the data. The variables that are finally present in the resultant trees are used in the next step of the analysis.

#### 3.3.2 Multiple regression

Multiple regression is used to explain the relationship between one continuous dependent variable with two or more independent variables. It is an extension of simple linear regression. The result of the multiple regression indicates whether the outcome or dependent variable can be predicted from the independent variables, the accuracy of the prediction, model fits and statistical significance. Multiple regression has several assumptions on the data that should be tested to find whether the method is suitable for the dataset. These assumptions [18] are – dependent variables should be continuous and linearly dependent on independent variables, at least two or more independent variables – which can be either continuous or categorical and should not be highly correlated, independence among the observations, homoscedasticity in the data and normally-distributed residuals.

In this research, we generated three multiple regression models for each of the dependent variables – social distance, personal protection behaviour, social responsibility awareness. After applying CART in the previous step, three independent variables are excluded from the overall nine independent variables. Thus, each of the three multiple regression models has six independent variables – age, sex, marital status, child, drinking frequency and smoking frequency.

## 4. Results

Table 2 provides a summary of the research dataset of this study. As notable, the dataset contained a balance of male and female respondents and different demographic attributes. Table 3 shows the corresponding RIDIT scores for the five responses against each of 21 activities. The survey respondents reported these activities in response to a survey question (i.e., “*Have you ever conducted anything to prevent novel coronavirus infections or outbreaks?*”). The responses of these activities were considered to quantify the three dependent variables (S*ocial distance, Personal protection behaviour, Social responsibility awareness*) of this study.

**Table 2:** Basic statistics of the research dataset.

|  | **All** | **Female** | **Male** |
| --- | --- | --- | --- |
| *Total respondents* | 11,342 | 5,608 | 5,734 |
| *Age (years)* |  |  |  |
| 1. 20-29 | 1,964 (17.32) | 970 (17.30) | 994 (17.34) |
| 2. 30-39 | 2,336 (20.60) | 1,140 (20.33) | 1,196 (20.86) |
| 3. 40-49 | 3,098 (27.31) | 1,530 (27.28) | 1,568 (27.35) |
| 4. 50-59 | 2,754 (24.28) | 1,381 (24.63) | 1,373 (23.94) |
| 5. 60-64 | 1,190 (10.49) | 587 (10.47) | 603 (10.52) |
| *Marital status* |  |  |  |
| 1. Married | 4,722 (41.63) | 2,229 (39.75) | 2,493 (43.48) |
| 2. Not married | 6,620 (58.37) | 3,379 (60.25) | 3,241 (56.52) |
| *Having children younger than junior high school age* |  |  |  |
| 1. No | 2,972 (26.20) | 1,479 (26.38) | 1,493 (26.04) |
| 2. Yes | 8,370 (73.80) | 4,129 (73.62) | 4,241 (73.96) |
| *Drinking frequency* |  |  |  |
| 1. Never drink | 3,980 (35.09) | 2,360 (42.08) | 1,620 (28.25) |
| 2. I used to drink, but I quit | 860 (7.58) | 518 (9.24) | 342 (5.96) |
| 3. Few times per month | 2,216 (19.54) | 1,142 (20.36) | 1,074 (18.73) |
| 4. 1-2 times per week | 1,640 (14.46) | 697 (12.43) | 943 (16.45) |
| 5. 3-6 times per week | 1,139 (10.04) | 388 (6.92) | 751 (13.10) |
| 6. Every day | 1,507 (13.29) | 503 (8.97) | 1,004 (17.51) |
| *Smoking frequency* |  |  |  |
| 1. Never smoked | 6,748 (59.50) | 4,120 (73.47) | 2,628 (45.83) |
| 2. Used to smoke but not now | 2,188 (19.29) | 773 (13.78) | 1,415 (24.68) |
| 3. Sometimes | 270 (2.38) | 82 (1.46) | 188 (3.28) |
| 4. Everyday | 2,136 (18.83) | 633 (11.29) | 1,503 (26.21) |
| *Work status* |  |  |  |
| 1. Regular employee | 5,817 (51.29) | 1,831 (32.65) | 3,986 (69.52) |
| 2. Non-regular employee | 2,865 (25.26) | 2,132 (38.02) | 733 (12.78) |
| 3. Self-employed and others | 660 (5.82) | 238 (4.24) | 422 (7.36) |
| 4. Not working | 2,000 (17.63) | 1,407 (25.09) | 593 (10.34) |

**Table 3:** RIDIT score for the five responses against each of 21 activities of the survey.

| Response | Frequency | RIDIT score |
| --- | --- | --- |
| Not at all | 16146 | 0.07 |
| Not true | 28152 | 0.19 |
| Neither | 57141 | 0.43 |
| True | 75809 | 0.74 |
| Very true | 60934 | 1.00 |

Figures 2–4 illustrate the corresponding CART outcomes for the three independent variables of this study. We used SPSS [19] with default settings for generating these three trees. Three attributes (*Age, Income* and *Education*) were not found in each of three CART models for the three dependent variables – an indication that these variables may have low influence on the dependent variables. For this reason, these three variables were not considered in the multi regression model considered in this research.

**Figure 2:**
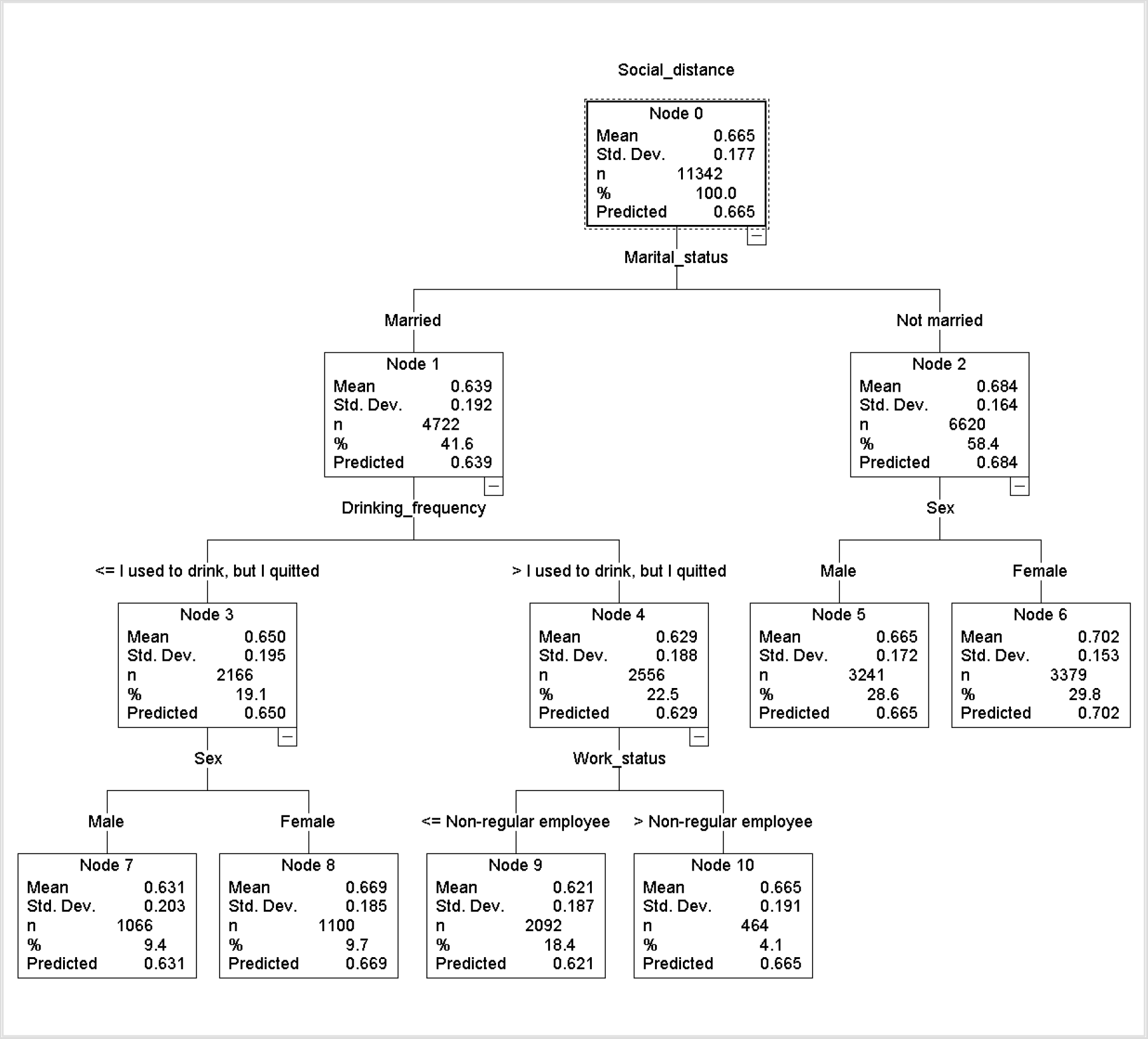
CART outcome tree for the ‘*Social distance*’ dependent variable. Out of nine, four attributes (*Marital status, Drinking frequency, Sex* and *Work status*) were included as nodes in this tree.

**Figure 3:**
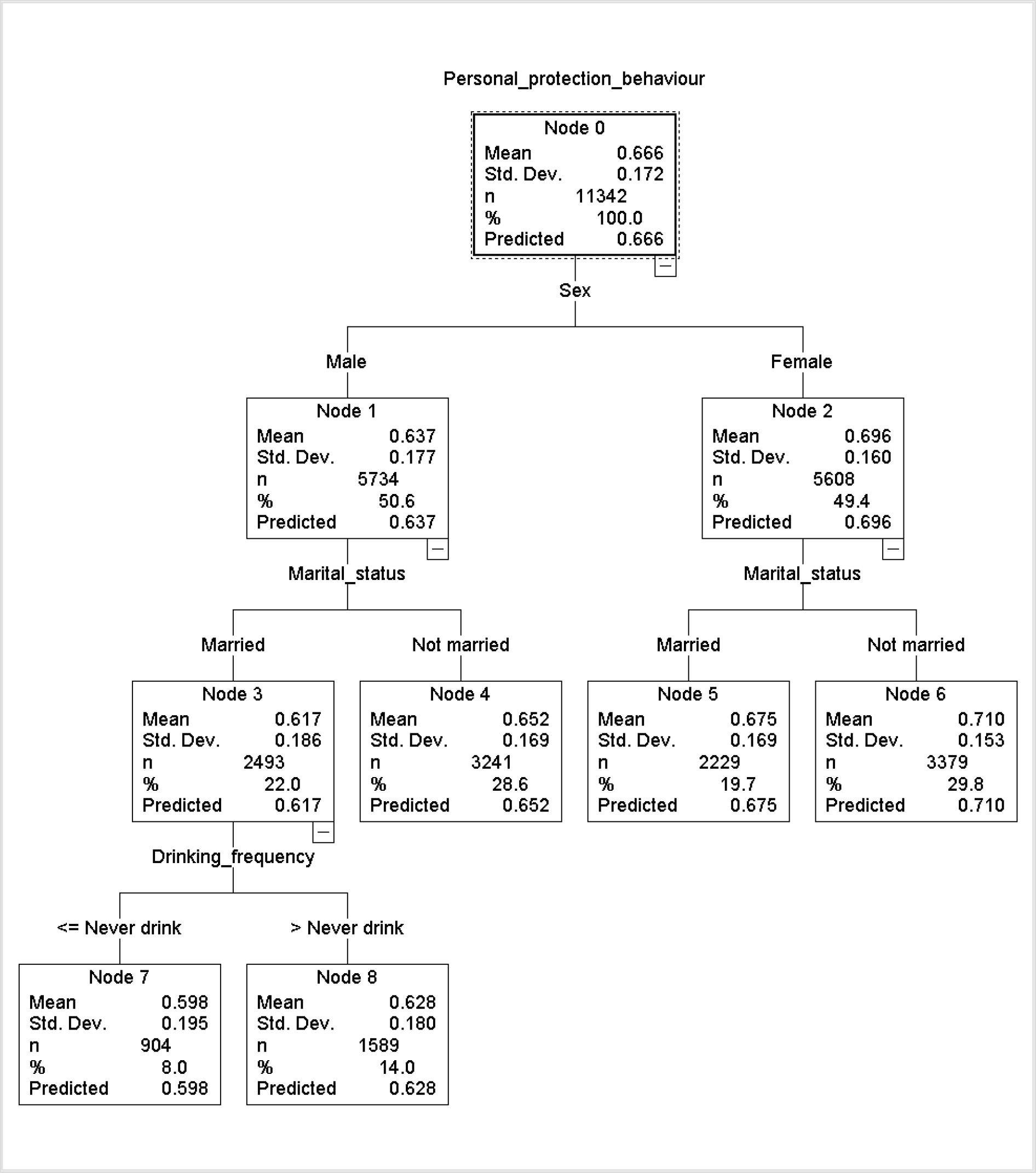
CART outcome tree for the ‘*Personal protection behaviour*’ dependent variable. Out of nine, four attributes (*Sex, Marital status* and *Drinking frequency*) were included as nodes in this tree.

**Figure 4:**
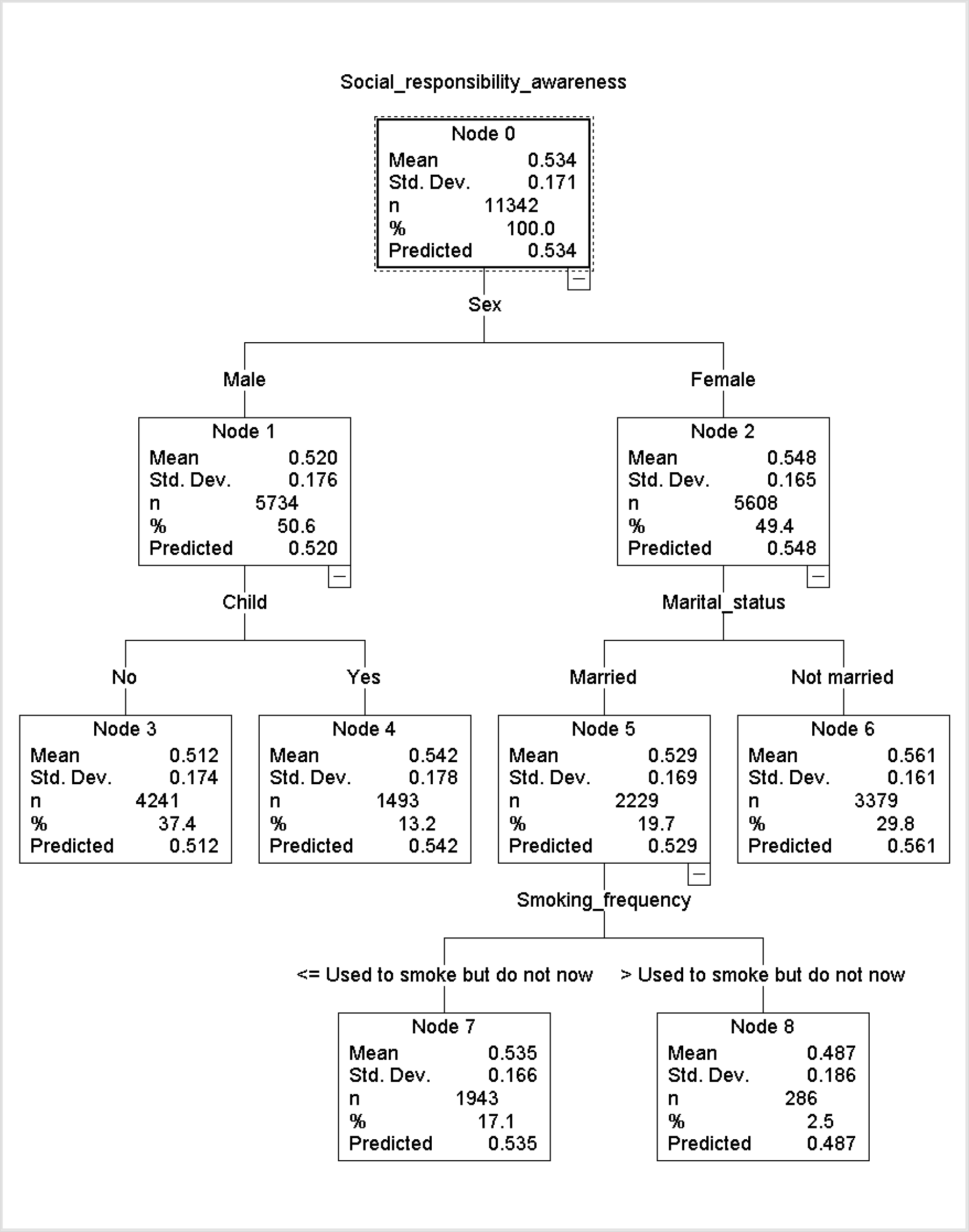
CART outcome tree for the ‘*Social responsibility awareness*’ dependent variable. Out of nine, five attributes (*Sex, Child, Marital status* and *Smoking frequency*) were included as nodes in this tree.

Table 4 shows the three regression models developed for each of three dependent variables of this study. Three attributes (*Child, Marital status* and *Sex*) have significant positive impact on each of the three dependent variables (*Social distance, Personal protection behaviour* and *Social responsibility awareness*). The ‘*drinking frequency*’ attribute does not have a significant impact on any of the dependent variables. The ‘*smoking frequency*’ has negative impact on all dependent variables but this impact is not significant for the ‘*personal protection behaviour*’ variable. The ‘*work status*’ attribute has significant positive impact on the ‘*social distance*’ variable but significant negative impact on the ‘*personal protection behaviour*’ variable. It does not have a significant impact on the ‘*social responsibility awareness*’ variable. We also considered VIF (variance inflation factor), which is a statistical measure that can detect the presence of multicollinearity in a regression analysis. In regression analyses, multicollinearity is a phenomenon which is used to explore the presence of any linear relations among predictor variables [19]. In general, a VIF value of > 10 indicates a high presence of multicollinearity. As revealed by the VIF (variance inflation factor) values indicated in Table 4, multicollinearity is not present in our regression analysis.

**Table 4:** Regression outcome for the three dependent variables considered in this study

|  | Model One<br>Dependent variable: *Social distance* |  |  |  | Model Two<br>Dependent variable: *Personal protection behaviour* |  |  |  | Model Three<br>Dependent variable: *Social responsibility awareness* |  |  |  |
| --- | --- | --- | --- | --- | --- | --- | --- | --- | --- | --- | --- | --- |
|  | Beta | Sig. | VIF | R<sup>2</sup> | Beta | Sig. | VIF | R<sup>2</sup> | Beta | Sig. | VIF | R<sup>2</sup> |
| (Constant) | 0.591 | 0.000 |  | 0.028 | 0.564 | 0.000 |  | 0.042 | 0.516 | 0.000 |  | 0.016 |
| Child | 0.034 | 0.001 | 1.219 |  | 0.038 | 0.000 | 1.219 |  | 0.046 | 0.000 | 1.219 |  |
| Drinking frequency | -0.016 | 0.104 | 1.108 |  | 0.012 | 0.204 | 1.108 |  | 0.007 | 0.469 | 1.108 |  |
| Marital status | 0.110 | 0.000 | 1.231 |  | 0.084 | 0.000 | 1.231 |  | 0.050 | 0.000 | 1.231 |  |
| Sex | 0.057 | 0.000 | 1.170 |  | 0.174 | 0.000 | 1.170 |  | 0.064 | 0.000 | 1.170 |  |
| Smoking frequency | -0.030 | 0.002 | 1.112 |  | -0.018 | 0.057 | 1.112 |  | -0.059 | 0.000 | 1.112 |  |
| Work status | 0.059 | .0000 | 1.108 |  | -0.022 | 0.023 | 1.108 |  | 0.010 | 0.324 | 1.108 |  |

As revealed in Table 4, females are more likely to follow COVID-19 preventive practices than males. During the regression analyses, the numerical values of ‘1’ and ‘2’ are used for males and females, respectively. So, a positive coefficient in Table 4 for the ‘*sex*’ attribute indicates a higher adherence of females in following the preventive practices. As mentioned earlier, the ‘*child*’ attribute has also a positive impact on each of the three preventive measures of this study. We further categorise the survey respondents based on their ‘*sex*’ and ‘*child*’ information and, using *t-test*, explore the difference in abiding by the COVID-19 preventive practices among those categories. As outlined in Table 5, regardless of having a child, females tend to adhere preventive practices more than males.

**Table 5:** *t-test* results to explore the differences in abiding by the COVID-19 preventive practices between different subgroups of the survey respondents.

| Test no. | Independent variable | Group | N | Mean | STD | t-value | Sig. |
| --- | --- | --- | --- | --- | --- | --- | --- |
| 1 | Social distance | Female having child | 4,129 | 0.670 | 0.172 | 6.702 | 0.000 |
|  |  | Male having child | 4,241 | 0.644 | 0.187 |  |  |
| 2 |  | Female not having child | 1,479 | 0.711 | 0.154 | 7.521 | 0.000 |
|  |  | Male not having child | 1,493 | 0.665 | 0.178 |  |  |
| 3 | Personal protection behaviour | Female having child | 4,129 | 0.690 | 0.162 | 16.645 | 0.000 |
|  |  | Male having child | 4,241 | 0.628 | 0.178 |  |  |
| 4 |  | Female not having child | 1,479 | 0.712 | 0.156 | 8.675 | 0.000 |
|  |  | Male not having child | 1,493 | 0.661 | 0.174 |  |  |
| 5 | Social responsibility awareness | Female having child | 4,129 | 0.543 | 0.165 | 8.226 | 0.000 |
|  |  | Male having child | 4,241 | 0.512 | 0.174 |  |  |
| 6 |  | Female not having child | 1,479 | 0.563 | 0.166 | 3.419 | 0.001 |
|  |  | Male not having child | 1,493 | 0.542 | 0.178 |  |  |

## 5. Discussion

Overall, among the demographic attributes and personal habits (i.e. smoking, drinking behaviour) considered for this research, it appears that the demographic attributes like sex, marital and family status have played an influential role in abiding by COVID-19 protection behaviours in Japan. Research notes that, during the H1N1 pandemic, sex and marital status differently characterised intentions to get vaccinated among the Italian health workers [20]. Another research highlights that males were more adoptive of protection behaviours during the H1N1 pandemic in Saudi Arabia [21], while females were noted as more compliant to H1N1 prevention measures in Hong Kong [22]. A research further notes that the reason for vaccination following the H1N1 pandemic varied between males and females in Japan [23]. The COVID-19 outbreak, although a different disease, has some similarities to the past pandemic and is also caused by a virus. The findings from this study therefore reaffirm the differences in pandemic protection behaviour in population due to demographic attributes like sex. Further, research highlights that women tended to show more infection protection behaviour than men across countries during the previous pandemics [24]. Thus, this study’s finding of Japanese women being more compliant to COVID-19 protection measures matches this attitude. The analysis also reveals ‘*work status*’ as a contributing factor concerning two perspectives – ‘*social distancing*’ and ‘*personal protection behaviour*’. Some research has noted work status of individuals having an impact on their compliance to pandemic protection measure [24], and our finding thus affirms work status as a socio-demographic factor to consider during pandemic management.

Further to the socio-demographic attributes, this research focused on personal attributes including smoking and drinking frequency. While *‘drinking frequency’* appears non-significant for the considered models, the ‘*smoking frequency*’ attribute reveals a negative impact on each of the three COVID-19 preventive measures of this study. This indicates that a lower value of this measure is more desirable since such values will lead to a higher possibility of following preventive practices. As outlined in Table 2, the possible values for the ‘*smoking frequency*’ attribute have been placed in ascending order (i.e., ‘1’ for ‘*never smoked*’, ‘2’ for ‘*used to smoked but not now*’, ‘3’ for ‘*sometimes*’ and ‘4’ for ‘*everyday*’). Thus, a negative coefficient for the ‘*smoking frequency*’ attribute indicates that individuals who are non-smoker or smoke less frequently are more likely to adhere COVID-19 preventive measures. Thus, a negative coefficient for the ‘*smoking frequency*’ attribute relates to non-smoking or infrequent smoking behaviour, and which is apparently linked to more adherence to COVID-19 preventive measures. Research notes that smoking and non-smoking behaviour can characterise the practice of seeking health support for influenza types of diseases in the USA [25]. Another study finds smoking behaviour as a contributing factor towards vaccination against H1N1 among pregnant women in France [26]. Thus, this study’s finding of ‘*smoking behaviour*’ of Japanese citizens shaping their adherence to COVID-19 restrictions affirms this personality attribute as a factor to consider in pandemic management.

This research considered three constructs (*social distancing, personal protection* and *social responsibility awareness*) that are commonly suggested and followed across the world to mitigate the spread of COVID-19. In quantifying these three constructs, the 21 different responses of a single survey question were used. These 21 responses were grouped into three equal-sized (i.e., seven) groups for these three constructs. These groupings are statistically reliable, as revealed in the corresponding Cronbach’s alpha values of Table 6. The seven responses of each group are closely related to each other. Cronbach’s alpha is a measure of internal consistency and reliability used to explore ‘*how closely a set of items are related*’ when they are used to quantify a construct [27]. In general, a value of 0.70 or more is considered as ‘acceptable’ in most social science research situations.

**Table 6:**
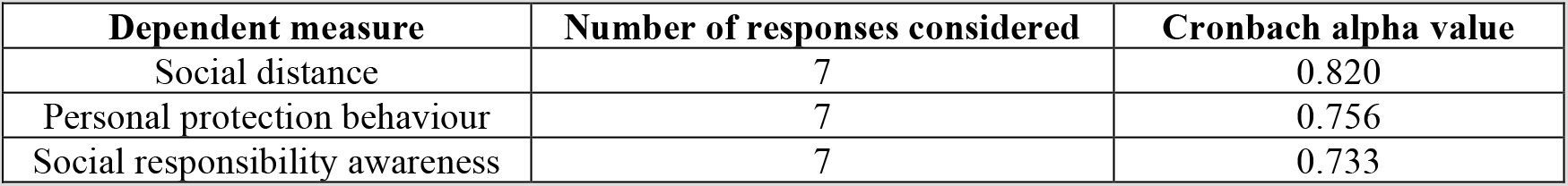
Cronbach’s alpha score for each of the three dependent measures considered in this study.

Both ‘*child*’ and ‘*marital status*’ have significant positive impact on each of the three COVID-19 preventive measures considered in this study. This indicates individuals who have a younger child and are not married are more likely to adhere these preventive practices. In our research dataset, on average, married individuals (1.95) have more younger children than ‘*not married*’ individuals (1.58). The ‘*not married*’ status includes both ‘*unmarried*’ and ‘*married but divorced*’ individuals. Since only a small percentage of Japanese children are born to unmarried mothers [28], most of the ‘*not married*’ respondents in the research data are more likely to be divorced individuals. In recent years, divorce tendency has become more common in the USA and many other developed countries including Japan [29]. So, our findings related to ‘*child*’ and ‘*marital status*’ should be considered carefully in developing COVID-19 preventive practice policies for countries where single parenting is historically low due to cultural and religious reasons (e.g., India).

Finally, it is worth noting the technical contribution made in this research. As outlined earlier, while a recently published article on the same dataset also explores Japanese populaces’ response to pandemic protection measures and advice [13], we zoom into the pandemic protection measures from three behavioural perspectives and considered more items of the respective survey question. In addition, rather than opting only for descriptive statistics and multiple linear regression, we use the CART analysis to extract features that are influential in characterising the dependent variables. Further, CART can conceptualise non-parametric and nonlinear relationship among data [30], and can therefore avoid limitations in a linear regression. Our results, thus, complement the existing finding [13], and our approach may also motivate similar interpretative categorisation of pandemic protection behaviour in future research. Indeed, with socio-demographic and personal attributes differently impacting the different protection behaviours, a policy implication can be instituting a targeted policy response across different aspects of pandemic protection measures for the different socio-demographic groups.

## 6. Conclusion

This study assesses the behavioural perspectives of population differentiated by socio-demographic and personal attributes when confronting a pandemic situation. Using a publicly available and recently collected data on Japanese citizens during the COVID-19 early outbreak and exploiting both CART and regression analysis, the study notes that socio-demographic and personal attributes of individuals indeed shape the subjective prevention actions and thereby the control of spread of a pandemic. Three socio-demographic attributes – sex, marital family status, and having children – appear to have played an influential role in Japanese citizens’ abiding by the COVID-19 protection behaviours, especially with women having children being noted more conscious than the male counterparts. Among the personality attributes, smoking behaviour appeared as a contributing factor with non-smokers or less-frequent smokers more compliant to the protection behaviours. Work status also appears to have some impact especially concerning social distancing and personal protection behaviour.

There are some limitations of the research. The conclusions drawn are dependent on information recorded in the publicly available dataset. Thus, the study may have sustained the same limitations as the data collector including limitations due to self-reporting by participants, not covering all age groups, random sampling, and timing of data collection [13]. Further, the way the items for the considered questionnaire have been interpreted and grouped across the three behaviours can be subjective, as is not uncommon in qualitative coding. Despite the limitations, the findings provide useful insights especially concerning the need for public policy campaigning to account for variations in responses and protection behaviour due to socio-demographic and personal attributes during a pandemic. Such consideration, rather than a general campaign, may lead to more effective pandemic management – an issue to be explored in future research.

## Data Availability

This research used data that are publicly available

## Author statements

### Ethical approval

This research does not need any ethical approval since it uses a publicly available dataset for research analysis purposes.

## References

1. Liu, Y., Gayle, A.A., Wilder-Smith, A., and Rocklöv, J., The reproductive number of COVID-19 is higher compared to SARS coronavirus. J Travel Med, 2020. 27(2).

2. Kannan, S., Ali, P.S.S., Sheeza, A., and Hemalatha, K., COVID-19 (Novel Coronavirus 2019)-recent trends. Eur. Rev. Med. Pharmacol. Sci, 2020. 24(4): p. 2006–2011.

3. Mustafa, N.M. and A Selim, L., Characterisation of COVID-19 Pandemic in Paediatric Age Group: A Systematic Review and Meta-Analysis. Journal of clinical virology: the official publication of the Pan American Society for Clinical Virology, 2020. 128: p. 104395–104395.

4. Setti, L., Passarini, F., De Gennaro, G., Barbieri, P., Perrone, M.G., Borelli, M., Palmisani, J., Di Gilio, A., Piscitelli, P., and Miani, A., Airborne transmission route of COVID-19: why 2 meters/6 feet of inter-personal distance could not Be enough. 2020. 17(8): p. 2932.

5. World Health Organisation. Coronavirus disease (COVID-2019) situation reports. 2020; Available from: https://www.who.int/emergencies/diseases/novel-coronavirus-2019/situation-reports.

6. Djalante, R., et al., Review and analysis of current responses to COVID-19 in Indonesia: Period of January to March 2020. Progress in Disaster Science, 2020. 6: p. 100091–100091.

7. Zhan, S., Yang, Y.Y., and Fu, C., Public’s early response to the novel coronavirus-infected pneumonia. Emerging microbes & infections, 2020. 9(1): p. 534–534.

8. Shabu, S., Amen, K., Mahmood, K., and Shabila, N., Risk perception and behavioral response to COVID-19 in Iraqi Kurdistan Region. 2020.

9. Wadood, M.A., Lee, L.L., Huq, M., Mamun, A., Mohd, S., and Hossain, M., Practice and perception of Bangladeshi adults toward COVID-19: a cross-sectional study. 2020.

10. Lai, A.Y.-H. and Tan, S.L., Impact of Disasters and Disaster Risk Management in Singapore: A Case Study of Singapore’s Experience in Fighting the SARS Epidemic, in Resilience and Recovery in Asian Disasters: Community Ties, Market Mechanisms, and Governance, D.P. Aldrich, S. Oum, and Y. Sawada, Editors.2015 Springer Japan: Tokyo. p. 309–336.

11. Fakhruddin, B., Blanchard, K., and Ragupathy, D., Are we there yet? The transition from response to recovery for the COVID-19 pandemic. Progress in Disaster Science, 2020. 7: p. 100102.

12. Yamamoto, I., Japanese citizens’ behavioral changes. Ann Arbor, MI: Inter-university Consortium for Political and Social Research (distributor),2020

13. Muto, K., Yamamoto, I., Nagasu, M., Tanaka, M., and Wada, K., Japanese citizens’ behavioral changes and preparedness against COVID-19: An online survey during the early phase of the pandemic. PLOS ONE, 2020. 15(6): p. e0234292.

14. Bross, I.D., How to use ridit analysis. Biometrics, 1958: p. 18–38.

15. D. Flora Jr, J., Ridit analysis. Wiley StatsRef: Statistics Reference Online, 2014.

16. Bross, I.D., How to use ridit analysis. Biometrics, 1958. 14(1): p. 18–38.

17. Breiman, L., Friedman, J., Stone, C.J., and Olshen, R.A., Classification and regression trees. 1984: CRC press.

18. Laerd Statistics. Multiple regression analysis using SPSS statistics. Lund Research Ltd 2013 [cited 2020 21-06-2020]; Available from: https://statistics.laerd.com/spss-tutorials/multiple-regression-using-spss-statistics.php.

19. Field, A., Discovering statistics using SPSS. 2009: Sage Publications Ltd.

20. La Torre, G., Di Thiene, D., Cadeddu, C., Ricciardi, W., and Boccia, A., Behaviours regarding preventive measures against pandemic H1N1 influenza among Italian healthcare workers, October 2009. Eurosurveillance, 2009. 14(49): p. 19432.

21. Balkhy, H.H., El-Saed, A., and Sallah, M., Epidemiology of H1N1 (2009) influenza among healthcare workers in a tertiary care center in Saudi Arabia: a 6-month surveillance study. infection control and hospital epidemiology, 2010. 31(10): p. 1004–1010.

22. Lau, J.T., Griffiths, S., Choi, K.C., and Tsui, H.Y., Avoidance behaviors and negative psychological responses in the general population in the initial stage of the H1N1 pandemic in Hong Kong. BMC Infectious Diseases, 2010. 10(1): p. 139.

23. Iwasa, T. and Wada, K., Reasons for and against receiving influenza vaccination in a working age population in Japan: a national cross-sectional study. BMC public health, 2013. 13(1): p. 647.

24. Bish, A. and Michie, S., Demographic and attitudinal determinants of protective behaviours during a pandemic: A review. British journal of health psychology, 2010. 15(4): p. 797–824.

25. Biggerstaff, M., Jhung, M.A., Reed, C., Garg, S., Balluz, L., Fry, A.M., and Finelli, L., Impact of medical and behavioural factors on influenza-like illness, healthcare-seeking, and antiviral treatment during the 2009 H1N1 pandemic: USA, 2009–2010. Epidemiology & Infection, 2014. 142(1): p. 114–125.

26. Freund, R., Ray, C.L., Charlier, C., Avenell, C., Truster, V., Tréluyer, J.-M., Skalli, D., Ville, Y., Goffinet, F., Launay, O., and Group, f.t.I.C.S., Determinants of Non-Vaccination against Pandemic 2009 H1N1 Influenza in Pregnant Women: A Prospective Cohort Study. PLOS ONE, 2011. 6(6): p. e20900.

27. Cronbach, L.J., Coefficient alpha and the internal structure of tests. psychometrika, 1951. 16(3): p. 297–334.

28. Raymo, J.M., Single motherhood and children’s health and school performance in Japan. Marriage & family review, 2016. 52(1–2): p. 64–88.

29. Ogihara, Y., The rise in individualism in Japan: Temporal changes in family structure, 1947–2015. Journal of Cross-Cultural Psychology, 2018. 49(8): p. 1219–1226.

30. vanEngelsdorp, D., et al., Weighing Risk Factors Associated With Bee Colony Collapse Disorder by Classification and Regression Tree Analysis. Journal of Economic Entomology, 2010. 103(5): p. 1517–1523.

